# Ultrasonic Artificial Intelligence Shows Statistically Equivalent Performance for Thyroid Nodule Diagnosis to Fine Needle Aspiration Cytopathology and *BRAFV600E* Mutation Analysis Combined

**DOI:** 10.1101/2022.04.28.22274306

**Authors:** Tianhan Zhou, Lei Xu, Jingjing Shi, Yu Zhang, Tao Hu, Rujun Xu, Lesi Xie, Lijuan Sun, Dandan Li, Wenhua Zhang, Chuanghua Chen, Wei Wang, Chenke Xu, Fanlei Kong, Yanping Xun, Lingying Yu, Shirong Zhang, Jinwang Ding, Fan Wu, Tian Tang, Siqi Zhan, Jiaoping Zhang, Dexing Kong, Dingcun Luo

## Abstract

**Objective:** To investigate the difference between an artificial intelligence (AI) system, fine-needle aspiration (FNA) cytopathology, *BRAFV600E* mutation analysis and combined method of the latter two in thyroid nodule diagnosis.

**Methods:** Ultrasound images of 490 thyroid nodules (378 patients) with postsurgical pathology or twice of consistent combined FNA examination outcomes with a half-year interval, which were considered as gold standard, were collected and analyzed. The diagnostic efficacies of an AI diagnostic system and FNA-based methods were evaluated in terms of sensitivity, specificity, accuracy, *κ* coefficient compared to the gold standard.

**Results:** The malignancy threshold of 0.53 for an AI system was selected according to the optimization of Youden index based on a retrospective cohort of 346 nodules and then applied for a prospective cohort of 144 nodules. The combined method of FNA cytopathology according to Bethesda risk stratification system and *BRAFV600E* mutation analysis showed no significant difference in comparison with the AI diagnostic system in accuracy for both the retrospective and prospective cohort in our single center study. Besides, for the 33 indeterministic Bethesda system category III and IV nodules included in our study, the AI system showed no significant difference in comparison with the *BRAFV600E* mutation analysis.

**Conclusion:** The evaluated AI diagnostic system showed similar diagnostic performance to FNA cytopathology combined with and *BRAFV600E* mutation analysis. Given its advantages in ease of operation, time efficiency, and noninvasiveness for thyroid nodule screening as well as the wide availability of ultrasonography, it can be widely applied in all levels of hospitals and clinics to assist radiologists for thyroid nodule diagnosis and is expected to reduce the need for relatively invasive FNA biopsies and thereby reducing the associated risks and side effects as well as to shorten the diagnostic time.

---

Thyroid nodules are a common endocrine disease. Approximately 5% of adults have palpable thyroid nodules, and incidental thyroid nodules are detected in 19-67% of ultrasound examinations, most of which are asymptomatic^1,2^. The key to thyroid nodule evaluation is to distinguish benign from malignant thyroid nodules^3^. Thyroid Imaging Reporting & Data Systems (TI-RADS) have been widely adopted by radiologists in the world for thyroid nodule risk stratifications^4^. The diagnosis accuracy by radiologists according to TI-RADS standards are highly varying, depending on their personal experiences and subjective judgement. The Bethesda system for reporting thyroid cytopathology based on minimally invasive Fine Needle Aspiration (FNA), can effectively distinguish the majority of benign and malignant nodules, providing effective guidance for clinical thyroid nodule management. Pathological studies show that the Bethesda I-VI diagnostic categories correspond to the 5-10%, 0-3%, 10-30%, 25-40%, 50-75% and 97-99% of malignancy risks, suggesting that even for most certain categories II and VI, there can still exist misdiagnoses. More importantly, two categories, i.e., III and IV respectively known as “atypical or follicular lesion of undetermined significance” and “follicular neoplasm/suspicious for follicular (or Hürthle cell) neoplasm”, are unreliable for estimating the real malignancy risk, resulting in 15-30% nodules with inconclusive cytological results^5^.

It has been shown that genetic tests can help improve the cytological diagnostic accuracy for thyroid nodules, among which a specific biomarker for Papillary Thyroid Carcinoma (PTC), namely *BRAFV600E*, has been widely applied in the diagnosis of thyroid nodules^6^. In western countries, the mutation rate of *BRAFV600E* is about 40-45%^7^, while in Asian people, its mutation rate is about 52-83%, significantly higher than in the west^8^. It has been found that the *BRAFV600E* mutation analysis is especially effective for discriminating thyroid nodules belonging to the Bethesda III category^9^. It has also been found that combining the diagnosis of cytopathology and genetic testing can improve diagnostic accuracy^10^. However, it consumes more time and raises the overall cost. Furthermore, it may demand more FNA samplings, causing more unpleasant experiences to patients.

In recent years, with the introduction of the concept of precise diagnosis and treatment, and the progress of science and technology, the application of artificial intelligence (AI) in the medical field has developed rapidly. The diagnostic efficacy of AI for thyroid nodules has been shown to have comparable to or even better than that of senior radiologists^11-13^. In the clinical workflow of thyroid nodule risk assessment, thyroid nodule evaluation by radiologists is followed by FNA-based cytopathology and then by postsurgical pathology, which is the gold standard when necessary. Currently, there has been no study comparing the diagnostic efficacies of AI-assisted diagnosis and FNA-based cytopathology and BRAF V600E mutation combined with gold standard as the final diagnosis.

An ultrasonic AI-assisted diagnostic system, AI-SONIC™ Thyroid, has been increasingly equipped in Chinese hospitals of different levels ranging from the first-tier research-oriented ones to the local communal clinics for assisting radiologists for thyroid nodule risk evaluation. It spins from a university research project with the first publications dated back in 2017^14,15^ and is continuously evolving with active algorithm development and an increasing dataset collected from all over China and labeled with specifically designed review strategies by radiologists. It is built based on the EfficientNet architecture^16^ using the proprietary lightweight C++ deep learning framework DE-Light which is about 10% of Caffe in code length. The neural network is initialized using self-training with noisy student method on the ImageNet database. A focal loss function has been defined to resolve the problem of unbalanced sample distribution and increase the learning weights of difficult samples. Besides, a sharpness perception minimization algorithm is used to reduce both the loss and the loss sharpness to improve the generalization ability of the model. It is of general interest to assess the clinical value of this ultrasonic AI-assisted diagnostic system in comparison with FNA for discriminating benign and malignant thyroid nodules, taking the postsurgical pathology or twice of consistent combined FNA examination outcomes with a half-year interval as the diagnostic standard.

For optimizing the diagnostic performance of the AI system, the choice of the cut-off value for malignancy risk probability was very critical. This optimal cut-off value for the AI system was chosen according to the Youden index^17^ based on the cohort of retrospectively collected 997 ultrasound images of 346 thyroid nodules. We then compared the diagnostic performances of the AI system to that of the FNA cytopathology and genetic testing. Thereafter, we collected another 441 ultrasound images of 144 thyroid nodules prospectively with the objective to verify the results of the retrospective study.

## 1. Data and methods

### 1.1 Research subjects

In this study, the clinical data of patients with thyroid nodules admitted to Affiliated Hangzhou First People’s Hospital, Zhejiang University School of Medicine from July 2017 to August 2020 were collected and retrospectively analyzed, and those of patients with thyroid nodules admitted from September 2020 to December 2021 were collected and prospectively analyzed. Inclusion criteria: 1) patients aged 18-75 years, without gender preference, 2) those whose nodules had complete and standard sectional views observed horizontally and vertically, 3) those who had received FNA for thyroid nodules, and 4) those whose nodules were confirmed by postsurgical pathological examinations or twice of consistent combined FNA examination outcomes with a half-year interval. Exclusion criteria: 1) patients with nodules <2 mm or >50 mm, 2) those with uncontrolled subacute thyroiditis or hyperthyroidism, or 3) those who had previously received thyroid surgery or radiofrequency ablation of thyroid nodules. This study was approved by the ethics committee of our hospital. The inclusion and exclusion criteria are illustrated in Figure 1.

**Figure 1:**
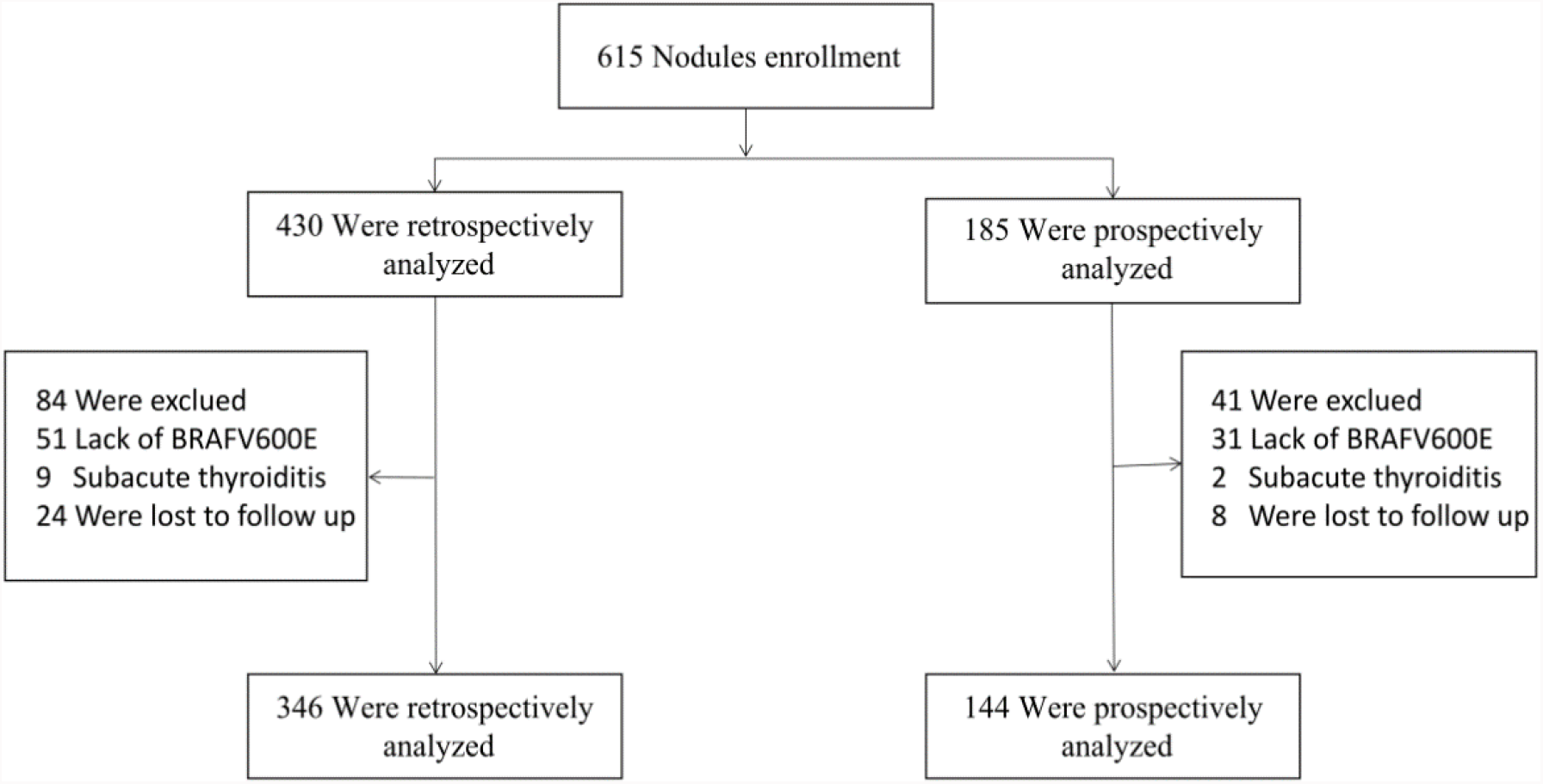
Flowchart of inclusion and exclusion criteria.

### 1.2 Equipment, instruments, and AI-assisted diagnostic system

ESAOTE color Doppler ultrasound machine equipped with a real-time, high-frequency (5-10 MHz) linear-array probe was used in this study. Ultrasound images of all nodules were collected and stored according to the protocol specified in the guideline^4^.

The computer-aided diagnostic system evaluated in this study, namely AI-SONIC™ Thyroid (version 5.3.0.2), is developed by Demetics Medical Technology Ltd (Hangzhou, China). It takes individual B-mode thyroid ultrasound images as input, detects each nodule automatically by displaying a bounding-box and computes the malignancy score associated with the nodule, with a value ranging from 0 to 1 as the output. It also allows interactive segmentation by radiologists as an additional input accompanying the original B-mode image in case the automatically detected nodule is not satisfactory, based on which the malignancy score is recomputed.

### 1.3 FNA of thyroid nodules

The FNA of thyroid nodules followed the *Revised American Thyroid Association management guidelines for patients with thyroid nodules and differentiated thyroid cancer (2009 Edition)*^*18*^. Aspiration without negative pressure was applied to target nodules using a special biopsy needle (23 G×50 mm, Hakko Corp., Japan). Each patient was placed in the supine position with the neck hyperextended and supported by a pillow under the shoulder, with the head leaning slightly towards the unaffected side to fully expose the site for aspiration. After routine disinfection, thyroid nodules were probed by ultrasound to determine the needle insertion paths. The ultrasound probe was held by the operator with one hand, and the biopsy needle was inserted obliquely along the scanning plane with the other hand, allowing real-time observation of the whole process. The needle insertion was stopped until the needle tip reached the center of the nodule, and then the plunger was pulled and pushed 5 times along different needle tracks. The aspirated specimens were smeared evenly on a slide and immediately fixed in 95% methanol for 10 min.

### 1.4 Cytological diagnosis of thyroid nodules

In this study, the Bethesda System for Reporting Thyroid Cytopathology *(2017 Edition)* was adopted^5^. According to the guidelines of Bethesda system on clinical management of categories I to VI thyroid nodules and practical need for binary risk classification, we decided to assign Bethesda categories I to IV to “negative cytology”, while categories V and VI were classified as “positive cytology”, which is consistent with other studies^10,19^.

### 1.5 Diagnosis by *BRAFV600E*

The nuclear acids were first isolated and purified from the FNA samples using AmoyDx tissue DNA/RNA kit ADx-TI03 (Amoy Diagnostics, Xiamen, China) according to the protocol provided by the manufacture. Ensuring the OD260/OD280 ratio of being within 1.8∼2.0, the purified nuclear acid concentrations were then determined by the NanoDrop™ 2000/2000c Spectrophotometer (Cat. No: ND-2000, Thermo Fisher Scientific), and were finally diluted to the concentration of 2.0ng/μL.

For FNA specimens, *BRAFV600E* point mutations were detected by amplification-refractory mutation system–PCR using the human *BRAFV600E* mutation detection kit ADx-BR01 (Amoy Diagnostics, Xiamen, China). 5 μL of the standardized DNA templates (10 ng in total) prepared above were pipetted from each case to 35 μL of the reaction mixture containing *Taq* enzyme to prepare the PCR system (40 μL in total). In each PCR run, a quality control standard (STD) and a no-template control (NTC, self-provided ultra-pure water) were set. The PCR amplification was done in the 7900HT fast real-time fluorescence quantitative PCR system (Applied Biosystems) based on the following three stages: (reaction at 95°C for 5 min) × 1 cycle, (reaction at 95°C for 25 s, at 64°C for 20 s, and at 72°C for 20 s) × 15 cycles, and (reaction at 93°C for 25 s, at 60°C for 35 s, and at 72°C for 20 s) × 31 cycles. Fluorescence signals, including the internal control HEX™ (Cat:7900RAN, BIOplastics) signal and the external control fluorescein amidite (FAM, Cat:B-79480, BIOplastics) signal, were collected at 60°C, and the system automatically calculated the Ct value of each specimen after the reaction. The mutation status of *BRAFV600E* in specimens was determined according to the protocol of ADx-BR01. The FAM-Ct value ≥28 meant that there was no *BRAFV600E* mutation, while the FAM-Ct value <28 confirmed the existence of the *BRAFV600E* mutation.

### 1.6 Follow-up protocol

In this study, if the nodules went through surgeries, the pathological results were taken as the gold standard for diagnosis. If the nodules were considered to be benign in the first combined method of FNA cytopathology and *BRAFV600E* mutation analysis, after a comprehensive evaluation, they were reexamined by combined FNA-based methods at half a year after the first examination. If these nodules remained unchanged during the follow-up and were evaluated to be still benign, they were considered to be benign. If malignant results were obtained in the second round FNA cytopathological examinations, these nodules received further surgical treatment.

### 1.7 Statistical methods

The data were statistically analyzed with IBM SPSS 26.0 software. The patient data concerning the overall sum, the gender distribution, the number of malignant and benign nodules determined according to the postsurgical pathology and the follow-up protocol were reported with descriptive statistics. The ages and nodule sizes were instead described by their mean values and the standard deviations, while the classification results determined by the FNA cytopathology, genetic testing and the AI system were described by the absolute quantities and the corresponding percentages with respect to the true values given by the gold standard. The sensitivity, specificity, accuracy and *κ* coefficient of all test methods were statistically assessed. The larger the *κ* coefficient, the higher the consistency of the test method with the gold standard. Specifically, a *κ* coefficient ≥0.75 indicated high consistency; a *κ* coefficient in the range of 0.40-0.75 indicated intermediate consistency, and a *κ* coefficient <0.40 suggested low consistency. Group comparisons were done with χ^2^ test, and the Receiver Operating Characteristic (ROC) curves were analyzed. *P*<0.05 was considered statistically significant.

## 2. Results

### 2.1 Summary of patient and nodule statistics

In this study, a total of 490 thyroid nodules (178 benign and 312 malignant ones) were included, coming from 374 females and 104 males, with a male: female ratio of 1:3.6. These patients were aged 46.65±12.07 years, and the diameter of the nodules was 8.59±6.13 mm. The basic statistics of the patients and nodules were summarized in Table 1. Among the 490 nodules, 997 ultrasound images of 346 nodules were retrospectively collected, while 441 ultrasound images of 144 nodules were prospectively acquired for this study.

**Table 1:** Basic information of included patients

|  | Retrospective dataset (n=346) | Prospective dataset (n=144) |
| --- | --- | --- |
| **Gender (n=378)** |  |  |
| Female | 265 (78%) | 109 (78%) |
| Male | 74 (22%) | 30 (22%) |
| **Age (mean±std)** | 46.92±12.14 | 46.02±11.95 |
| **Nodule size (mm)** | 8.81±6.56 | 8.06±5.00 |
| **Pathological diagnosis** |  |  |
| Benign | 128 (40%) | 50 (35%) |
| Malignancy | 218 (60%) | 94 (65%) |
| **Bethesda classification** |  |  |
| I | 7 | 4 |
| II | 122 | 50 |
| III | 22 | 8 |
| IV | 3 | 0 |
| V | 77 | 28 |
| VI | 115 | 54 |
| **BRAFV600E** |  |  |
| Mutation | 161 (47%) | 78 (55%) |
| No mutation | 185 (53%) | 66 (45%) |

### 2.2 Selection the cut-off value for the AI diagnostic system based in the retrospective cohort

The cut-off value for the malignancy risk probability returned by the AI thyroid diagnostic system was optimized by maximizing the Youden index using the gold standard specified in the method section, based on the retrospective cohort of 346 thyroid nodules. At the optimized cut-off value of 0.53 (Figure 2), the AI diagnostic model had sensitivity, specificity, and accuracy of 96.33%, 89.06%, and 93.64%, respectively (Table 2).

**Table 2.**
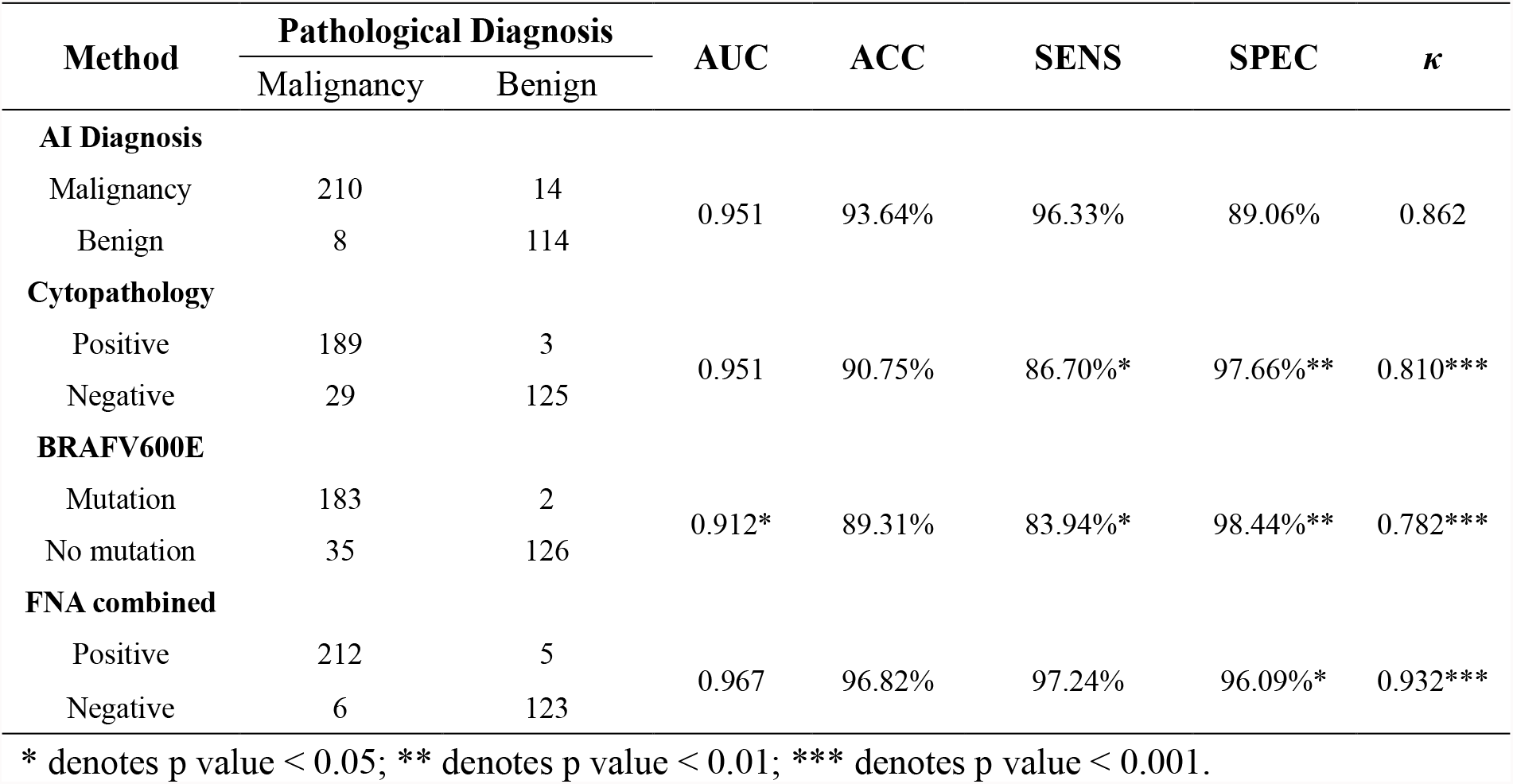
Diagnostic efficacies of different methods in retrospective cohort

| Method | Pathological Diagnosis | | AUC | ACC | SENS | SPEC | $\kappa$ |
| --- | --- | --- | --- | --- | --- | --- | --- |
|  | Malignancy | Benign |  |  |  |  |  |
| AI Diagnosis |  |  |  |  |  |  |  |
| Malignancy | 210 | 14 | 0.951 | 93.64% | 96.33% | 89.06% | 0.862 |
| Benign | 8 | 114 |  |  |  |  |  |
| Cytopathology |  |  |  |  |  |  |  |
| Positive | 189 | 3 | 0.951 | 90.75% | 86.70%* | 97.66%** | 0.810*** |
| Negative | 29 | 125 |  |  |  |  |  |
| BRAFV600E |  |  |  |  |  |  |  |
| Mutation | 183 | 2 | 0.912* | 89.31% | 83.94%* | 98.44%** | 0.782*** |
| No mutation | 35 | 126 |  |  |  |  |  |
| FNA combined |  |  |  |  |  |  |  |
| Positive | 212 | 5 | 0.967 | 96.82% | 97.24% | 96.09%* | 0.932*** |
| Negative | 6 | 123 |  |  |  |  |  |

\* denotes p value < 0.05; \*\* denotes p value < 0.01; \*\*\* denotes p value < 0.001.

**Figure 2:**
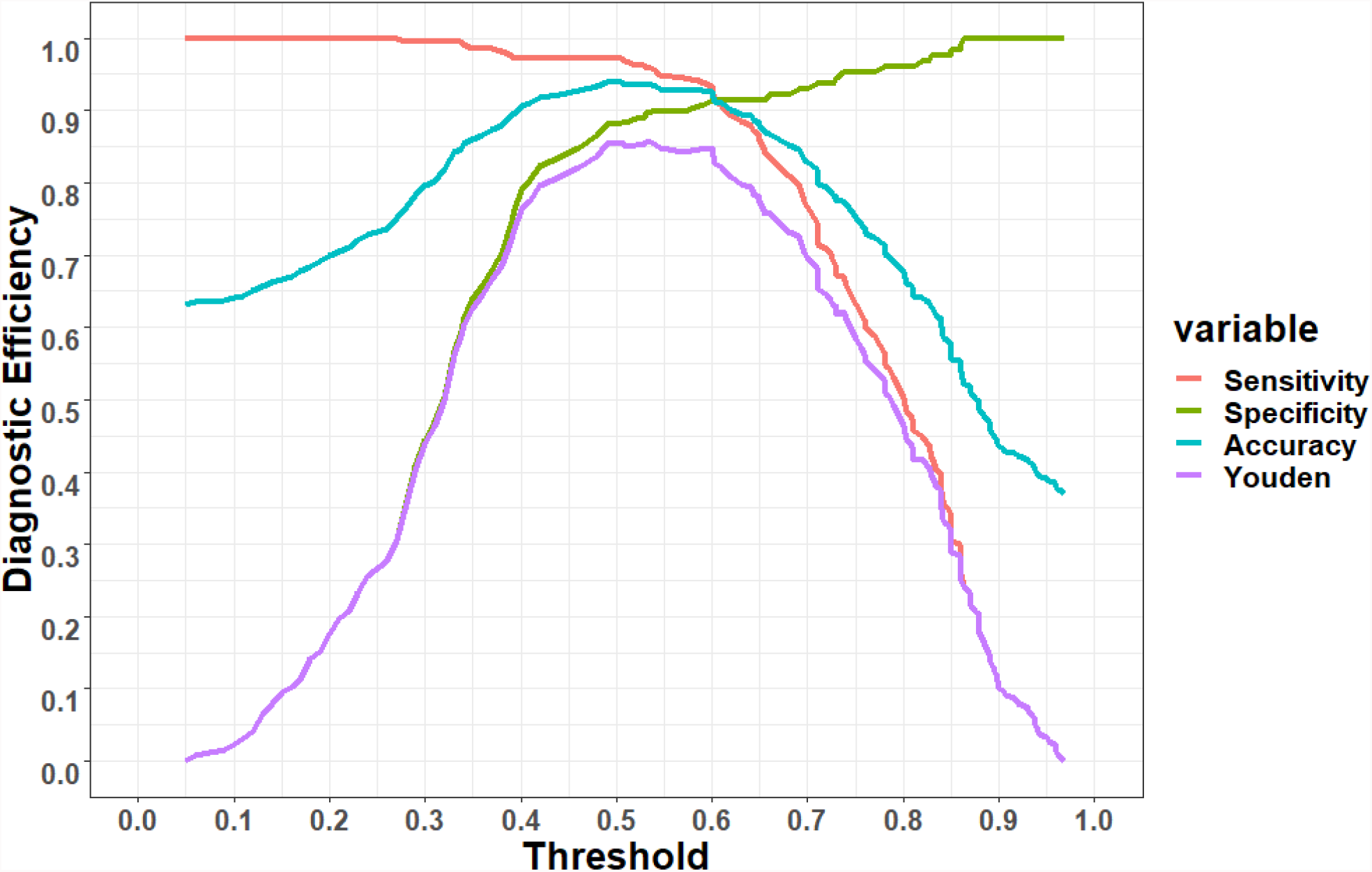
Selection of malignancy threshold for optimizing diagnostic efficacy of the AI system.

### 2.3 Diagnostic efficacy analysis of FNA on the retrospective cohort

#### 2.3.1 Diagnostic efficacy analysis of cytopathology

The FNA cytopathological examinations of the thyroid nodules were classified according to the Bethesda system. Categories I-IV were defined as negative cytological while categories V-VI represented positive results. In the retrospective cohort, 154 cases were considered negative and 192 cases positive by the cytopathological examinations. Its sensitivity, specificity, and accuracy in the malignancy diagnosis of thyroid nodules were 86.70%, 97.66%, and 90.75%, respectively (Table 2).

#### 2.3.2 Diagnostic efficacy analysis of *BRAFV600E*

In the retrospective cohort, *BRAFV600E* mutation was detected in 185 cases. In our study, no BRAFV600E mutation was classified as “negative”, while BRAFV600E mutation was classified as “positive”. The sensitivity, specificity, and accuracy of *BRAFV600E* mutation in diagnosing thyroid nodules were 83.94%, 98.44%, and 89.31%, respectively (Table 2).

#### 2.3.3 Diagnostic efficacy analysis of *BRAFV600E* combined with FNA cytopathology

We further diagnosed the thyroid nodules by combining the cytopathological examination with *BRAFV600E* mutation results. The diagnostic criterion was that thyroid nodules were considered as positive if either of the two examinations identified them as such, while the nodules were considered as negative when both methods suggested negative outcomes. In the retrospective cohort, the sensitivity, specificity, and accuracy of *BRAFV600E* mutation combined with FNA cytopathology in the diagnosis of thyroid nodules were 97.24%, 96.09%, and 96.82%, respectively (Table 2).

### 2.4 Comparison of diagnostic efficacy of the AI-assisted diagnostic system and FNA based methods

The ROC curves of each method were plotted and their corresponding values of the Area Under the Curve (AUC) were calculated to compare their diagnostic abilities for benign and malignant nodule discrimination (Figure 3). According to the consistency analysis using *κ* coefficient, there was a high consistency between each diagnostic method and the gold standard. However, the AI diagnostic system (0.862) had still significant higher *κ* coefficient than individual FNA-based methods, namely the FNA cytopathology (0.810) and *BRAFV600E* mutation analysis (0.782), but lower than their combined method (0.932) as shown in Table 2. In terms of overall diagnostic accuracy of thyroid nodules, the AI system did not significantly differ from the combined method (11/356 higher misdiagnosis rate with *P*-value=0.05), but it was significantly better than *BRAFV600E* mutation alone (*P*-value=0.017) and had 2.89% higher accuracy than the cytopathology alone (10/346), though without statistical significance (0.156) as one can see in Table 3. The ROC curves of different diagnostic methods in the retrospective cohort are shown in Figure 3. Moreover, for the retrospective cohort, the AI system showed significantly higher sensitivity than FNA-based methods, but the they showed higher specificity instead.

**Table 3:** Comparison of diagnostic efficacy between different methods in retrospective cohort

| Method | Diagnostic Accuracy | | $\chi^2$ | P-value |
| --- | --- | --- | --- | --- |
|  | Correct | Misdiagnosis |  |  |
| AI Diagnosis | 324 | 22 |  |  |
| Cytopathology | 314 | 32 | 2.009 | 0.156 |
| BRAFV600E | 309 | 37 | 5.740 | 0.017 |
| FNA combined | 335 | 11 | 3.061 | 0.050 |

**Figure 3:**
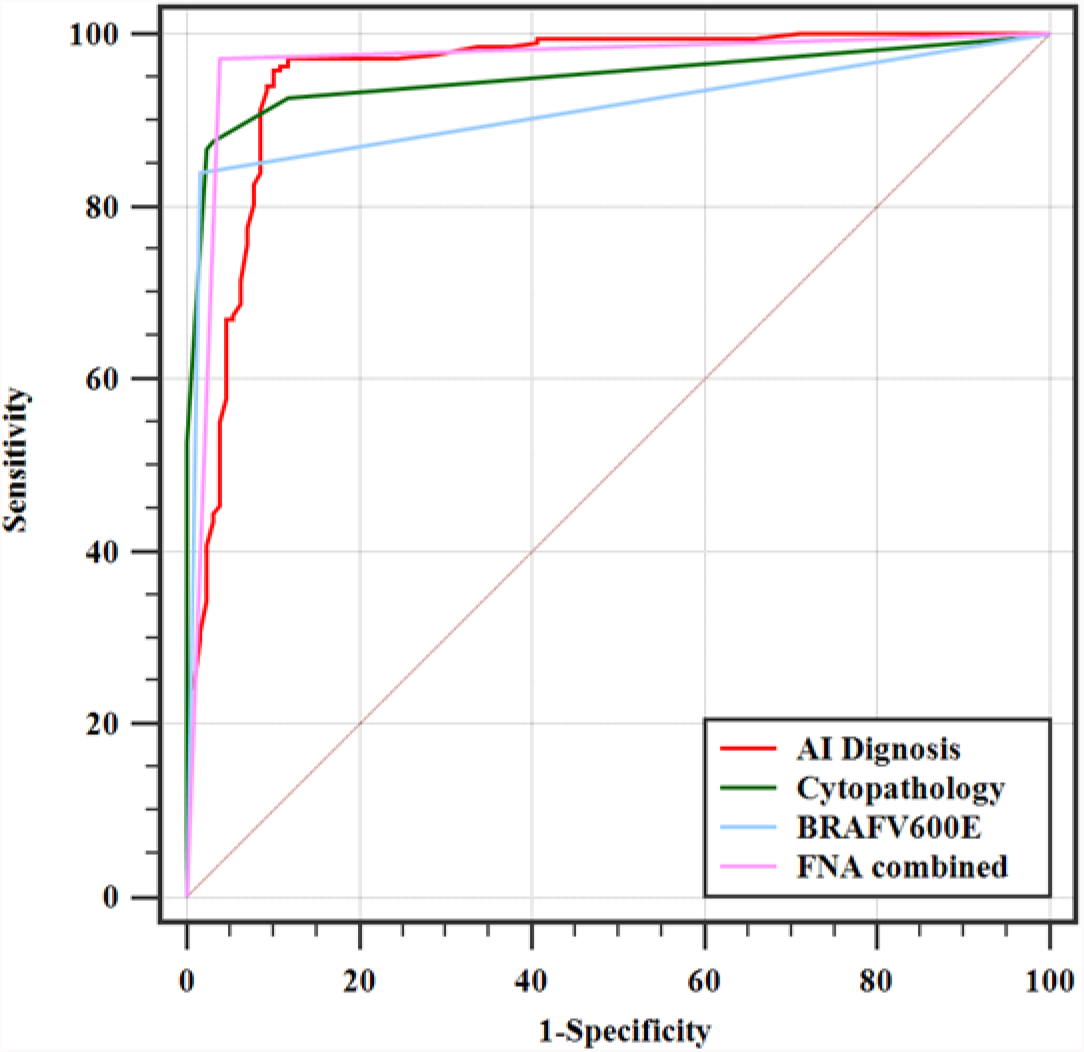
ROC curves of different diagnostic methods in the retrospective cohort

### 2.5 Diagnostic efficacies of different methods in prospective cohort and their statistical comparisons

In the cohort of prospective dataset of 144 nodules (Table 4), the sensitivity, specificity, and accuracy of the AI diagnostic system in identifying thyroid nodules using the same optimized cut-off value from the retrospective cohort were 93.62%, 94.00%, and 93.75%, respectively, while the sensitivity, specificity, and accuracy of FNA cytopathology in the diagnosis of benign and malignant thyroid nodules were 85.11%, 96.00%, and 88.89%, respectively. In contrast, the sensitivity, specificity, and accuracy of *BRAFV600E* mutation analysis in diagnosing thyroid nodules were 92.98%, 100%, and 88.89%, respectively. Furthermore, the sensitivity, specificity, and accuracy of the combined method in the diagnosis of thyroid nodules were 93.75%, 96.00%, and 94.44%, respectively. The AI diagnostic system had 4.86% (7/144) higher accuracy than both of the FNA cytopathology and *BRAF V600E* mutation analysis alone, though without statistical significance and had only one more misdiagnosis than their combined method (Table 5). Interestingly in the prospective cohort, in contrast to the retrospective cohort, the specificity of the AI system did not differ from individual FNA-based methods or their combination significantly. It can also be easily seen that for the prospective cohort, the ROC curve of the AI system had substantially larger area than the FNA cytopathology and *BRAFV600E* mutation analysis alone, and almost overlapped with that of their combined method, shown in Figure 4. However, due to limited prospective sample sizes, the differences in AUC values of all methods were considered insignificant.

**Table 4:** Diagnostic efficacies of different methods in the prospective cohort

| Method | Pathological Diagnosis | | AUC | ACC | SENS | SPEC | $\kappa$ |
| --- | --- | --- | --- | --- | --- | --- | --- |
|  | Malignancy | Benign |  |  |  |  |  |
| AI Diagnosis |  |  |  |  |  |  |  |
| Benign | 88 | 3 | 0.964 | 93.75% | 93.62% | 94.00% | 0.864 |
| Malignancy | 6 | 47 |  |  |  |  |  |
| Cytopathology |  |  |  |  |  |  |  |
| Positive | 80 | 2 | 0.909 | 88.89% | 85.11% | 96.00% | 0.768*** |
| Negative | 14 | 48 |  |  |  |  |  |
| BRAFV600E |  |  |  |  |  |  |  |
| Mutation | 78 | 0 | 0.915 | 88.89% | 92.98%* | 100.00% | 0.772*** |
| No mutation | 16 | 50 |  |  |  |  |  |
| FNA combined |  |  |  |  |  |  |  |
| Positive | 88 | 2 | 0.948 | 94.44% | 93.75% | 96.00% | 0.880*** |
| Negative | 6 | 48 |  |  |  |  |  |

\* denotes p value < 0.05; \*\*\* denotes p value < 0.001.

**Table 5:**
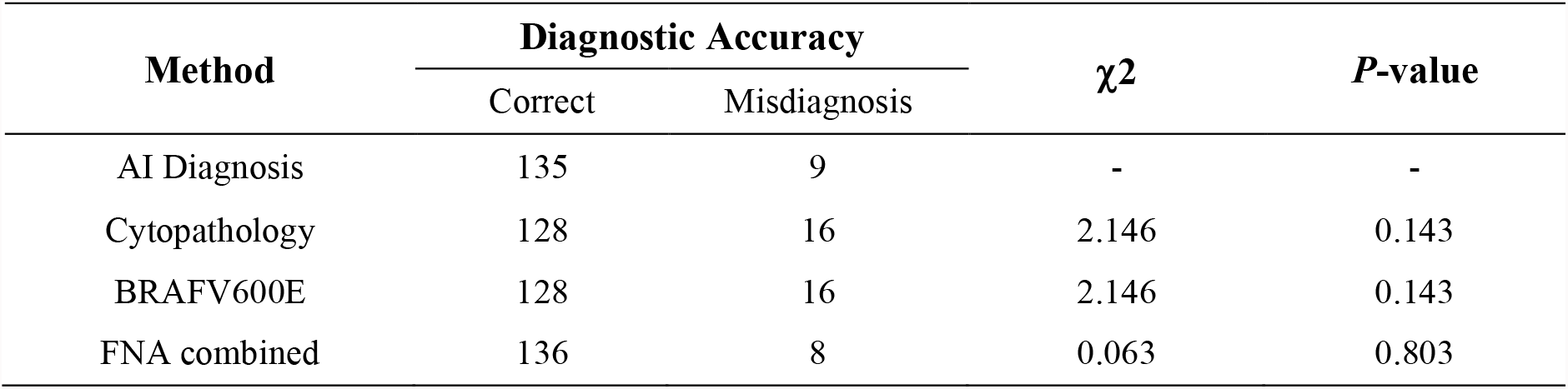
Comparison of diagnostic efficacy between different methods in the prospective cohort

**Figure 4:**
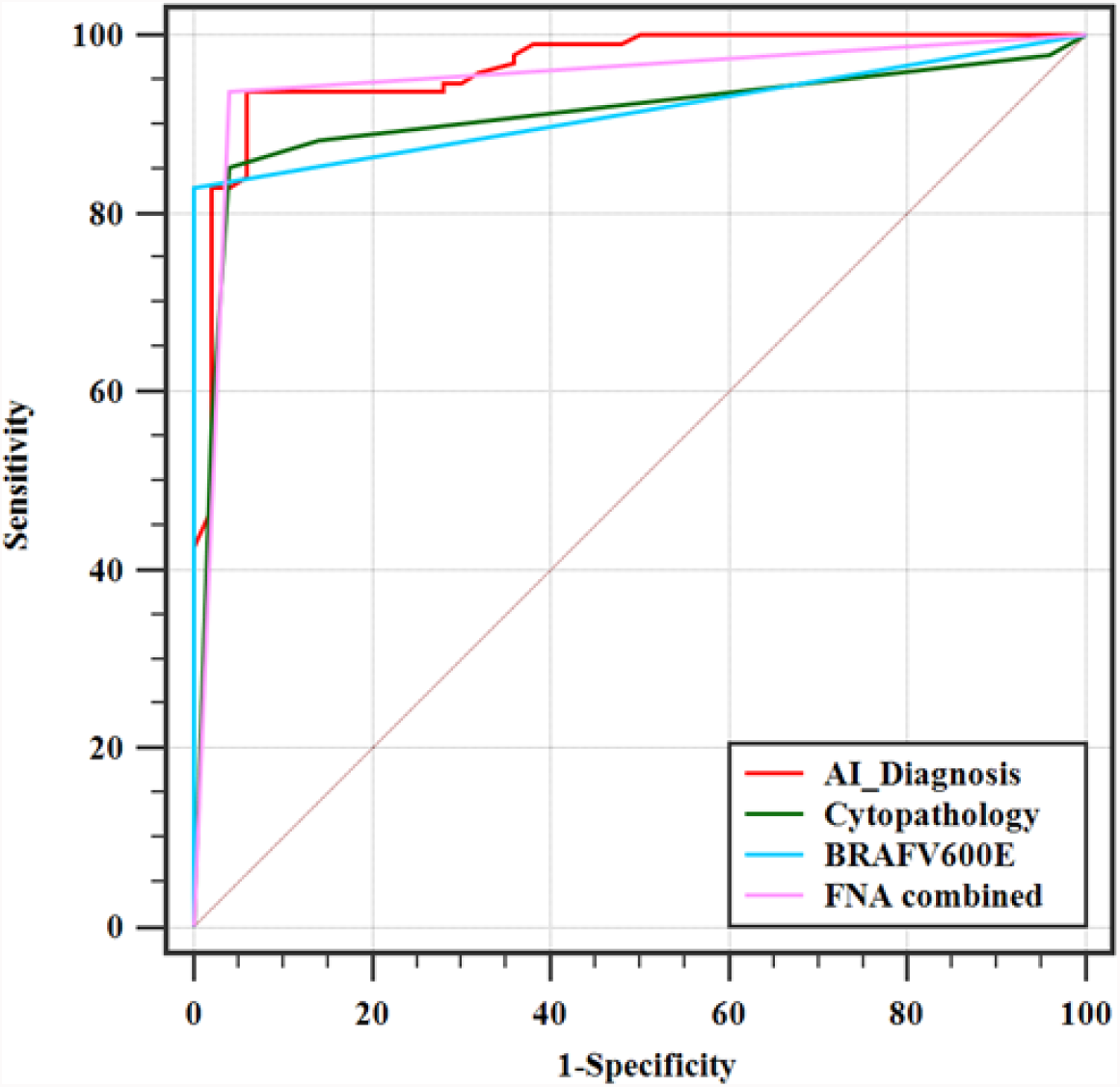
ROC curves of different diagnostic methods in the prospective cohort

### 2.6 Diagnostic evaluation of indeterminate thyroid nodules with Bethesda III and IV cytology

The enrolled nodules included 33 indeterminate thyroid nodules, consisting of 30 category III nodules and 3 category IV nodules, among which 16 were determined to be malignant and 17 were diagnosed to be benign by postsurgical pathology or reaffirming half-year follow-up examination combining both of the cytopathology and BRAFV600E mutation analysis. The sensitivity, specificity, and accuracy of *BRAFV600E* in the diagnosis of indeterminate thyroid nodules were 81.25%, 88.24%, and 84.85%, respectively. The sensitivity, specificity, and accuracy of the AI-assisted diagnostic system in diagnosing indeterminate thyroid nodules were 93.75%, 88.24% and 90.91% (Table 6). The difference in accuracy was not statistically significant due to limited number of samples. Representative images of a postsurgical pathological H&E staining, AI diagnosis, cytopathological staining as well as PCR curve of *BRAFV600E*, are shown in Figure 5 to illustrate the benefit of the AI system for these cases, where the Bethesda system for cytopathological classification provided indeterministic result, which however can be correctly predicted by the mutation analysis.

**Table 6:** Diagnostic efficacy of cytology for indeterminate thyroid nodules

| Method |  | Pathological diagnosis |  | SENS | SPEC | ACC | *P*-Value |
| --- | --- | --- | --- | --- | --- | --- | --- |
|  |  | Malignancy | Benign |  |  |  |  |
| BRAFV600E | Mutation | 13 | 2 | 81.25% | 88.24% | 84.85% | 0.708 |
|  | No Mutation | 3 | 15 |  |  |  |  |
| AI | Malignancy | 15 | 2 | 93.75% | 88.24% | 90.91% | - |
|  | Benign | 1 | 15 |  |  |  |  |

**Figure 5:**
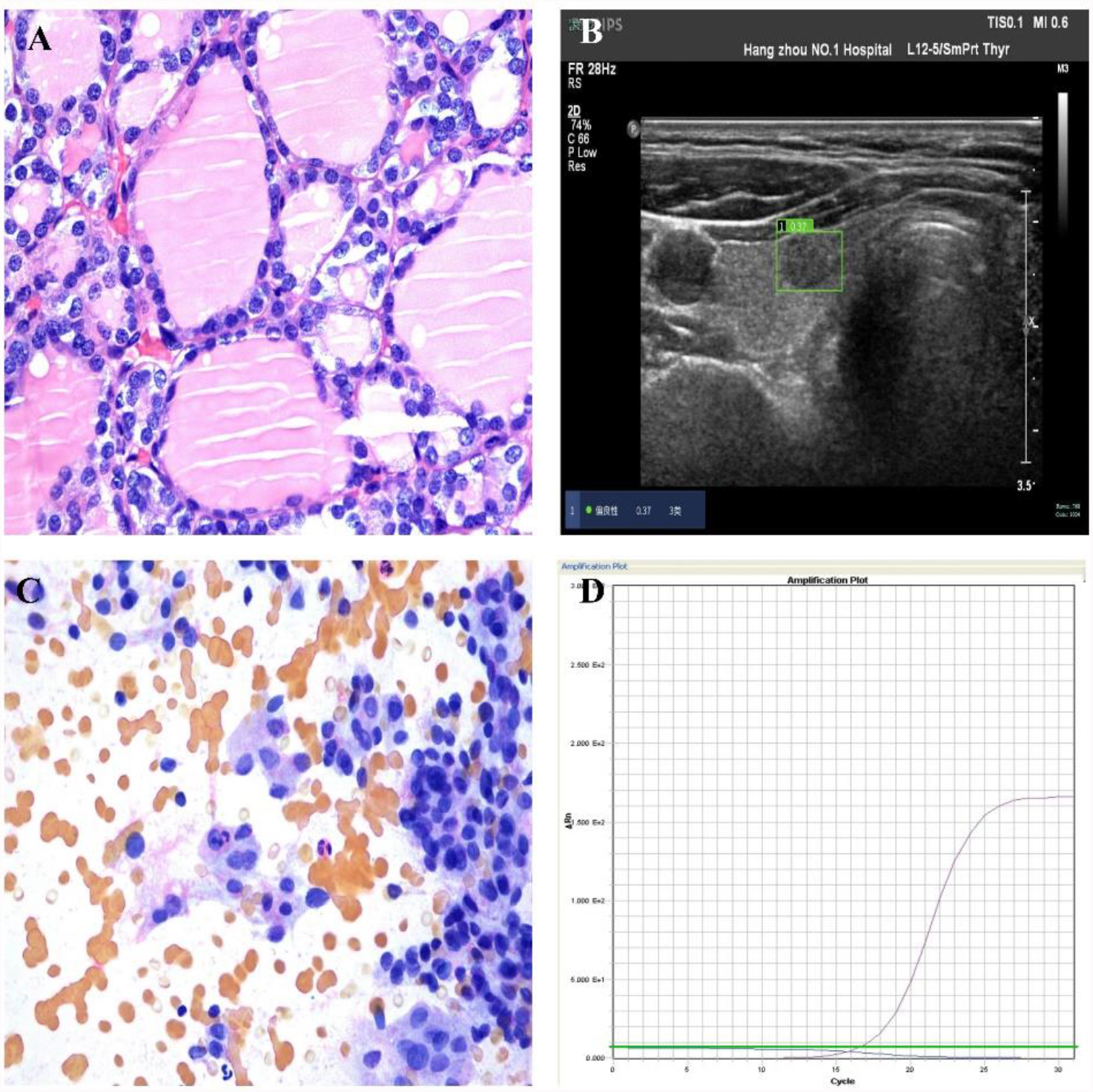
A presentative nodule of nodular goiter diagnosed by post-operative pathology (A) was correctly predicted by AI that returned a malignancy score of 0.37 (B), while FNA cytopathology reported uncertain classification but correctly predicted by *BRAF* analysis without mutation (D).

## 3. Discussion

Thyroid ultrasonography is the most common method used for screening of high-risk thyroid nodules, while the FNA-based cytopathology is the most common method for nodule diagnosis before surgical operation. Bethesda categories I to IV were classified as “negative cytology”, while categories V and VI were classified as “positive cytology” in our study, which is consistent with other studies^10,19^. As there were only 3 Bethesda category IV cases were included in our study, results would not be substantially affected regardless whether the category IV cases were counted as negative or positive cases. However, as the Bethesda risk stratification system for FNA cytopathology can get inconclusive results for 15-30% nodules, complementary genetic testing of the aspirated samples would be needed to improve the diagnostic efficacy of the thyroid nodules. *BRAFV600E* gene mutation was detected in 56.3% of the thyroid nodules (263/490). Results of the present study showed that the mutation rate of the malignant thyroid nodules was 83.65% (261/312), and benign thyroid nodules was 1.12% (2/178). Several publications demonstrated that the *BRAFV600E* gene was highly specific for diagnosing malignant nodules, but with low sensitivity, which is consistent with our study^20^. However, our represented data indicated a significantly higher sensitivity of BRAFV600E gene mutation than Rodrigues’s study^21^. It is likely related to the racial constitution, availability of puncture biopsy and different BRAFV600E gene mutation detection methods in each centre^6,7,22^. Three hundred and thirty-nine thyroid nodules underwent surgery and was confirmed by postsurgical pathological diagnosis in our study. And 151 thyroid nodules reexamined by combined method of FNA cytopathology and *BRAFV600E* mutation analysis after half a year follow-up was chosen.

In this study, we evaluated both the malignancy diagnostic efficacies of the FNA-based cytopathological examination and *BRAFV600E* mutation analysis in comparison to a deep learning-based AI automatic diagnostic system for thyroid nodules, taking the postsurgical pathological results or FNA twice as the gold standard. With an optimized cut-off value using the Youden index in the retrospective cohort of 346 thyroid nodules, the diagnostic accuracy of the AI system surpassed that of the *BRAFV600E* mutation analysis with statistical significance (*P*-value =0.017) and was statistically equivalent to FNA cytopathology examination (*P*-value = 0.156) and the combined method of FNA cytopathology with *BRAFV600E* testing (*P*-value = 0.05). Combining FNA cytopathology with *BFAF* improved the diagnostic accuracy of thyroid nodules from 90.75% to 96.82%, which is in line with the literature^19,23^. In the prospective cohort of 144 thyroid nodules, we showed that the AI diagnostic system did not significantly differ from *BRAFV600E* mutation analysis combined with FNA cytopathology. Taking the retrospective and prospective data together, misdiagnosis by the FNA combined method occurred in 19 patients, two of which had *BRAFV600E* mutations, manifested as nodular goiters by postoperative pathology, while the other 17 patients showed no *BRAFV600E* mutations. It can be concluded that false-negative and false-positive interpretations of *BRAFV600E* mutation have a non-negligible impact on the FNA combined method. The AI system misdiagnosed in total 31 nodules in this study. Hashimoto nodules, shriveled nodules, etc. can manifest similar ultrasonic to malignant nodules, which can mislead diagnoses by radiologists and AI systems^24^.

The head-to-head comparison of diagnostic results in this study detected no difference in the diagnostic efficacy between the AI-assisted diagnostic system and FNA combined method for thyroid nodules in both of the retrospective and prospective cohorts. The AI system had stable sensitivity, and were consistently higher than the *BRAF V600E* mutation analysis with statistical significance, but shows no statistical significance with FNA cytopathology examination or the combined method. Concerning its specificity, despite of being lower than individual FNA-based methods in the retrospective cohort with a value of 89.06%, the differences were no longer significant in the prospective cohort with a value of 94.00%. This suggests its specificity is not necessarily lower than FNA-based methods, but likely slightly more dependent on the sample distributions. In contrast, the sensitivity of *BRAFV600E* mutation analysis seemed to be more sample-dependent (83.94% vs 92.98%), while the FNA cytopathology had sensitivity consistently lower than 90% (86.70% vs 85.11%). It should be however noted that for determining benign nodules, substantial fractions of them were based on the gold standard set by the consistent results of twice FNA combined examinations with a half-year interval. In this study, only one out of 151 nodules had a changed outcome in the half-year follow-up FNA combined examination. This however does not mean that the first-round FNA combined method had merely 0.66% of error in benignity diagnosis for these nodules, as the extraordinarily consistent result could also be a reflection of methodological limitation in cases beyond its diagnostic capacity. Methodologically, it would be encouraged to exclude these cases where the FNA combined method itself was considered the gold standard for diagnostic efficacy evaluation. The drawback of excluding them is that the remained benign cases would be dominated by those requiring postsurgical pathological examinations and thus represent only a small fraction of real-world benign cases, complicating the interpretation of results. On the other hand, it was unethical to demand all supposedly benign nodules going through surgical pathological examinations for diagnosis confirmation. As a consequence, the definition of gold standard for benign nodules seemed to provide a privilege for the FNA combined method in comparison study. Interestingly, the AI system still managed to deliver statistically comparable overall diagnostic accuracy compared with the FNA combined method.

For indeterminate thyroid nodules defined as category III and IV according to Bethesda risk stratification system for FNA cytopathology, David *et al*. used a 112-gene classifier ThyroSeq V3 and reached 94% of sensitivity and 82% of specificity ^25^. In this study, *BRAF V600E* typing was used to diagnose indeterminate thyroid nodules with an accuracy of 84.85%. Though somewhat lower than that of the ThyroSeq V3, polygenic testing typically requires more aspiration biopsies and would consume more time and raise the overall cost. The accuracy, sensitivity and specificity of the AI diagnostic system for indeterminate thyroid nodules reached 90.91%, 93.75%, 88.24% respectively suggesting a potential superiority in the diagnosis of indeterminate thyroid nodules. However, the dataset of indeterminate thyroid nodules defined by Bethesda system included in this study was quite small in size (33 in total). A prospective study with a large dataset would be necessary for verifying the observation made in this study.

Other limitations to this study are summarized as follows: 1) In the real-world clinical evaluation, ultrasound-based diagnosis is confirmed by radiologists, not by an AI system alone. However, because of the black-box nature of deep learning algorithms, the predictions made by deep learning-based AI systems lacks interpretability and how they distinguish benign from malignant nodules are not transferable to radiologists, making how to objectively integrate the predictions by an AI system to radiologists’ workflow a crucial problem. Therefore, to what degree an AI system can help improve the diagnostic efficacy of radiologists and reduce unnecessary aspiration biopsies for patients should be further investigated. 2) This was a single-center study, despite of having achieved good outcomes in the prospective cohort. In the future, a multicenter study with a larger sample size would be needed to further verify the results.

Nevertheless, given no statistical significance in differences of diagnostic accuracies between the AI system and the combined method of FNA cytopathology and *BRAF V600E* mutation analysis, it is expected that the AI system can be widely applied in all levels of hospitals and clinics to assist radiologists, to reduce the need for relatively invasive FNA biopsies for thyroid nodule diagnosis thereby avoiding those patients’ discomfort during FNA biopsy processes and risks for biopsy-related side-effects or complications^26,27^, as well as to shorten the diagnostic time. The advancement of AI technologies will certainly have a tremendous impact on the diagnostic processes of thyroid nodules and expectedly many more diseases and transformative consensuses in disease management may be reached in the future.

## 4. Conclusion

For the malignancy diagnosis of thyroid nodules, the diagnostic efficacy of an existing AI automatic diagnostic system was statistically equivalent to that of FNA cytopathology combined with the *BRAFV600E* mutation analysis. With high diagnostic accuracy, noninvasiveness, and high user-friendliness, the AI automatic diagnostic system can be an indispensable tool for radiologists for thyroid nodule diagnosis and is expected to reduce the need for relatively invasive FNA biopsies and thereby reducing the associated risks and side effects as well as to shorten the diagnostic time.

## Data Availability

All data produced in the present study are available upon reasonable request to the authors

## Data Availability

All data produced in the present study are available upon reasonable request to the authors

